# Gender differences in PTSD severity and pain outcomes: baseline results from the LAMP trial

**DOI:** 10.1101/2023.10.13.23296998

**Authors:** JK Friedman, BC Taylor, E Hagel Campbell, K Allen, A Bangerter, M Branson, G Bronfort, C Calvert, LJS Cross, M Driscoll, R Evans, JE Ferguson, A Haley, S Hennessy, LA Meis, DJ Burgess

## Abstract

**Background:** Post-traumatic stress disorder (PTSD) and chronic pain are highly prevalent comorbid conditions. Veterans dually burdened by PTSD and chronic pain experience more severe outcomes compared to either disorder alone. Few studies have enrolled enough women Veterans to test gender differences in pain outcomes [catastrophizing, intensity, interference] by the severity of PTSD.

**Aim:** Examine gender differences in the association between PTSD symptoms and pain outcomes among Veterans enrolled in a chronic pain clinical trial.

**Methods:** Participants were 421 men and 386 women Veterans with chronic pain who provided complete data on PTSD symptoms and pain outcomes. We used hierarchical linear regression models to examine gender differences in pain outcomes by PTSD symptoms.

**Results:** Adjusted multivariable models indicated that PTSD symptoms were associated with higher levels of pain catastrophizing (0.57, 95% CI [0.51, 0.63]), pain intensity (0.30, 95% CI [0.24, 0.37]), and pain interference (0.46, 95% CI [0.39, 0.52]). No evidence suggesting differences in this association were found in either the crude or adjusted models (all interaction p-values<0.05).

**Conclusion:** These findings may reflect the underlying mutual maintenance of these conditions whereby the sensation of pain could trigger PTSD symptoms, particularly if the trauma and pain are associated with the same event. Clinical implications and opportunities testing relevant treatments that may benefit both chronic pain and PTSD are discussed.

## Background

Post-traumatic stress disorder (PTSD) and chronic pain are highly prevalent comorbid conditions that affect the health and well-being of U.S Veterans (1,2). Those dually burdened by PTSD and chronic pain may experience more severe psychiatric functional impairments and greater health care utilization than those with either disorder alone (3–5). Women Veterans, the fastest growing demographic of Veterans, experience disproportionally high rates of PTSD, 13%, compared to 6% of men Veterans, and between 11-78% experience chronic pain, compared to 9-50% of men Veterans (6–10). Despite the growing body of knowledge on comorbid PTSD and chronic pain, few studies have enrolled enough women Veterans to test gender differences in pain outcomes (pain catastrophizing, interference, and intensity) in relation to PTSD symptoms. Understanding the potential gender variation in the severity of chronic pain outcomes associated with PTSD symptoms has implications for developing gender-sensitive assessment tools and treatment modalities for Veterans with PTSD and chronic pain.

Chronic pain is defined as pain lasting longer than three consecutive months and is associated with decreased quality of life, increased medical costs, and increased substance use (11–14). In addition to the higher musculoskeletal pain reported by women Veterans, they are more likely to report moderate or severe chronic pain (15,16). At least one study found that women Veterans experienced greater pain intensity, decreased physical functioning, and more affective distress despite reporting better self-perceived health than men (17). Additionally, gender role expectations and societal norms may influence how individuals perceive and communicate their pain experiences, potentially contributing to reported disparities in clinical pain assessment, diagnosis, and offered treatment modalities (18–20). Finally, previous studies have revealed higher pain catastrophizing, interference and intensity levels in Veteran and civilian women compared to Veteran and civilian men (21,22).

PTSD is a mental health disorder that may develop after experiencing or witnessing a traumatic event and then subsequently developing a pattern of symptoms, including intrusion, avoidance, negative cognitions, and mood, or alterations in arousal and reactivity(23). It is associated with an increased risk of additional mental health diagnoses, although most people who experience traumatic events do not ultimately develop PTSD. Men Veterans report proportionally more combat-related exposures linked to PTSD symptoms (24). However, women Veterans who served in post-9/11 combat zones, such as Iraq and Afghanistan, report similar levels of perceived threat in the war zone as men (25,26). Women Veterans also report a higher incidence of trauma exposures across the life course, including adverse childhood experiences, military sexual trauma, and adult interpersonal violence, which may result in higher rates of PTSD compared to civilian women (16,27).

Comorbid chronic pain and PTSD frequently co-occur in civilian and Veteran populations and are thought to have a bidirectional and mutually reinforcing relationship (28,29). Chronic pain and PTSD are thought to share common underlying mechanisms. The shared vulnerability and mutual maintenance theories have been used to explain the co-morbidity of these conditions. The shared vulnerability theory posits that people with comorbid PTSD and chronic pain share common factors, such as anxiety sensitivity, attentional bias, autonomic arousal, and psychobiological dysregulation, predisposing them to one or both conditions (28). The mutual maintenance model posits that in addition to shared vulnerability factors (e.g., attentional biases, anxiety sensitivity), PTSD and chronic pain may exacerbate and reinforce each other (29). This bidirectional influence may result in the experience of pain-related exacerbation of PTSD symptoms by retriggering recollections of painful injuries sustained during the traumatic event. Conversely, elevated PTSD symptoms may activate the body’s stress response and the experience of increased pain. The co-morbidity of chronic pain and PTSD has been found in treatment-seeking samples of PTSD and chronic pain patients (1,28). This mechanism has been well studied in predominantly male Veteran (30) and civilian (31) samples, which may obscure important differences in the manifestation and severity. While the findings of studies using Veteran samples support the central tenant of the mutual maintenance model, namely the mutually reinforcing and exacerbating influence of PTSD on chronic pain outcomes, very few have had adequate numbers of men and women Veterans to identify differential effects by gender (32). The current study assesses whether gender differences exist in association between PTSD symptoms and pain outcomes in a comparative sample of men and women Veterans.

This manuscript reports the associations between probable PTSD symptoms and three dimensions of chronic pain outcomes: pain catastrophizing (negative thinking about one’s pain), pain intensity (how strong the pain feels), and pain interference (how one experiences pain, pain interference with daily activities), in a sample of U.S. Veterans diagnosed with a chronic pain condition. Specifically, we assessed gender differences in the association between probable PTSD and chronic pain outcomes. Based on the higher prevalence of both PTSD and chronic pain among women Veterans, the study team hypothesized that gender differences would be present across strata of probable PTSD and chronic pain outcomes. This study extends previous research on the associations between PTSD and chronic pain due to the unique gender composition of our data, which includes approximately equal numbers of men and women Veterans with chronic pain.

## Methods

### Study design and sample

This is a secondary analysis of Veterans randomized for participation in the Learning to Apply Mindfulness to Pain (LAMP) study (*n*=811). The LAMP study was a three-arm, multi-site, randomized pragmatic trial designed to assess the efficacy of multimodal delivery of two mindfulness-based interventions compared to usual care for Veterans with chronic pain at one of the three participating sites. Participants were recruited from the Minneapolis, Durham, and Greater Los Angeles Veterans Affairs Healthcare Systems. To be eligible, participants needed to have: 1) at least two qualifying pain diagnoses within the same pain category on at least two separate occasions at least 90 days apart during the previous two years documented in their electronic health record (EHR), 2) a pain duration of at least six months, a pain severity score of at least four during the previous week on a Likert scale of 0-10, and 3) access to a smartphone and the internet. Participants were excluded if they had: 1) new diagnoses of schizophrenia, bipolar disorder, major depressive disorder, current psychotic symptoms, suicidality, or other psychosis within the past 18 months (assessed by chart review); or 2) were currently enrolled in another pain study or a Mindfulness-Based Stress Reduction (MBSR) program (assessed by survey). Participants were recruited between November 4, 2020 and May 25, 2022. Participants provided verbal consent, which was recorded in the study tracking database by the study recruitment coordinator. Additional study details are described in the protocol paper (33).

Of the participants (*n*=1945) who met eligibility criteria, 1,737 completed the baseline survey, and 811 were randomized to one of the intervention arms. Due to an a priori study aim to examine gender differences, women were oversampled and comprised 48% of participants.

### Measures

#### Predictors

*PTSD symptoms* were assessed using the PTSD Checklist for *Diagnostic and Statistical Manual of Mental Disorders-Fifth Edition* (PCL-5) (34). The PCL-5 is a 20-item self-report measure that assesses the presence and severity of PTSD symptoms over the past month (range, 0-80). Respondents are asked to rate how bothered they have been by each of the 20 items on a five-point Likert scale. Response options for all items are: “Not at all” (0), “A little bit” (1), “Moderately” (2), “Quite a bit” (3), or “Extremely” (4). Items are summed with scores of 31 or higher, indicating probable PTSD and a need for additional clinical assessment or PTSD treatment (35). PCL-5 scores were standardized and parameterized as continuous in the analysis.

#### Outcomes

*Pain Catastrophizing* was assessed using the Pain Catastrophizing Scale (PCS) (36). The PCS is a 13-item self-report instrument that asks respondents to reflect on past painful experiences and indicate their thoughts and feelings in response to pain (range, 0-52). Response options for all items are: “Not at all” (0), “To a slight degree” (1), “To a moderate degree” (2), “To a great degree” (3), or “All the time” (4). Items are summed with scores of 30 or higher, representing a clinically significant level of pain catastrophizing (36). PCS scores were standardized and parameterized as continuous in analysis.

*Pain intensity and interference.* The Brief Pain Inventory (BPI) subscales were used to assess participant experiences of pain intensity (range, 0-10) and interference (range, 0-10) during the past week (37,38). Pain interference was assessed by seven items asking the extent to which pain interferes with general activity, mood, walking ability, normal work, relations with other persons, sleep, and enjoyment of life on an 11-point numeric rating scale from “Does not interfere” (0) to “Completely interferes” (10). Pain intensity was assessed by four items asking participants to rate their worst, least, average, and current pain severity for the past 1 month on an 11-point numeric rating scale from “No pain” (0) to “Pain as bad as you can imagine” (10). BPI subscale scores were standardized and parameterized as continuous in analysis (39).

#### Covariates

*Gender* was identified by self-report on the study screening questionnaire. Answer options included: man, woman, another gender, and decline to answer. Participants who identified as either “another gender” or “decline to answer” were then re-coded for analysis using their administrative birth-sex as reported in EHR.

*Sociodemographic* variables included self-identified racial identity (“White,” “African American or Black,” “American Indian/Alaskan Native,” “Asian,” Native Hawaiian/Pacific Islander,” and “Multiracial”); ethnicity (“Hispanic or Latino” and “Not Hispanic or Latino”), highest level of educational attainment (“High school or less,” “Some college,” “Bachelor’s degree,” and “Master’s degree or beyond”), and age. The patient facility where participants were recruited (Minneapolis, Durham, or Greater Los Angeles VA Medical Centers) was also included in adjusted models.

#### Analysis

First, we conducted descriptive analyses summarizing distributions of all variables overall and stratified by gender identity (Table 1). Crude and sociodemographic adjusted linear regression models were used to quantify the association between PTSD symptoms and each pain outcome (pain catastrophizing, intensity, and interference). We fit two sets of models (crude and adjusted) models to assess the association between PTSD symptoms and each pain outcome. We additionally assessed effect measure modification by gender (whether the association between PTSD symptoms and each pain outcome differed by gender identity) by adding an interaction term to each model. Due to differences in the scale range of all outcomes, all estimates were standardized to represent the number of standard deviations of change in each pain outcome associated with a one standard deviation change in the exposure variable, PTSD symptoms. Statistical significance was assessed at alpha level p<0.05. All statistical analyses were conducted using Stata 17 (College Station, TX).

**Table 1.** Demographic Characteristics.

| **Table 1. Demographic Characteristics** | **Full sample**<br>**(N=807)** | **Men**<br>**(N=421)** | **Women (N=386)** | **p-**<br>**value\*** |
| --- | --- | --- | --- | --- |
| Age (Mean, sd) | 55 (12.9) | 58 (12.6) | 50 (11.8) | $p < 0.05$ |
| *Racial Identity* | | | | $p < 0.05$ |
| African American/ Black | 207 (26%) | 70 (17%) | 137 (36%) |  |
| American Indian/ Alaskan Native | 9 (1%) | 5 (1%) | 4 (1%) |  |
| Asian | 6 (1%) | 3 (0.75%) | 3 (0.75%) |  |
| Multiracial | 35 (4%) | 15 (4%) | 20 (5%) |  |
| Native Hawaiian/ Pacific Islander | 1 (0.1%) | * | 1 (0.25%) |  |
| White | 545 (68%) | 326 (78%) | 219 (57%) |  |
| *Ethnicity* |  |  |  | p>0.05 |
| Hispanic or Latino | 51 (6%) | 21 (5%) | 30 (8%) |  |
| Not Hispanic or Latino | 755 (94%) | 400 (95%) | 355 (92%) |  |
| *Employment Status* |  |  |  | p<0.05 |
| Working now | 333 (41%) | 152 (36%) | 181 (47%) |  |
| Disabled | 174 (22%) | 86 (20%) | 88 (23%) |  |
| Retired | 203 (25%) | 142 (34%) | 61 (16%) |  |
| Other | 97 (12%) | 41 (10%) | 56 (15%) |  |
| *Income* |  |  |  | p>0.05 |
| Live comfortably | 247 (31%) | 133 (32%) | 114 (30%) |  |
| Meet your basic expenses with a little left over for extras | 343 (43%) | 189 (45%) | 154 (40%) |  |
| Just meet your basic expenses | 187 (23%) | 88 (21%) | 99 (26%) |  |
| Don't even have enough to meet basic expenses | 30 (4%) | 11 (3%) | 19 (5%) |  |
| *Education* |  |  |  | p<0.05 |
| High school or less | 53 (7%) | 36 (9%) | 17 (4%) |  |
| Some college | 353 (44%) | 196 (47%) | 157 (41%) |  |
| Bachelor's degree | 220 (27%) | 109 (26%) | 111 (29%) |  |
| Master's degree or more advance education | 181 (22%) | 80 (19%) | 101 (26%) |  |
| **Behavioral variables (Mean, SD)** |  |  |  |  |
| Brief Pain Inventory (BPI)- Interference (0-10) | 5.6 (2.0) | 5.4 (1.9) | 5.8 (2.0) | p<0.05 |
| Brief Pain Inventory (BPI)- Intensity (0-10) | 5.5 (1.5) | 5.4 (1.5) | 5.7 (1.6) | p<0.05 |
| Pain catastrophizing (PCS) (0-52) | 22.4 (11.3) | 21.3 (11.4) | 23.6 (11.1) | p<0.05 |
| PTSD (PCL-5) (0-80) | 27.3 (19.5) | 26.0 (19.3) | 28.7 (19.6) | p<0.05 |

\*ttest or chisq test; (scale range)

## Results

### Participant Characteristics

A total of 807 participants provided complete data on PTSD, pain catastrophizing, pain intensity, and pain interference on the baseline survey (*n=*807/811). Women in our sample were more likely to be employed, have four-year or more advanced degrees, be a member of a racially minoritized group, and be younger compared to men. Mean and standard deviations of scores for all outcome variables are also shown in Table 1. Women reported statistically significant higher mean scores for all pain outcomes and PTSD symptoms compared to men.

### PTSD symptoms and pain outcomes

Table 2 shows the standardized estimates, 95% confidence intervals (CI), and results from the regression models examining the association between PTSD symptoms and pain outcomes (catastrophizing, intensity, and interference). Results are shown for both the crude and the model adjusted for sociodemographic characteristics. Results from the adjusted models indicated that PTSD symptoms were associated with higher levels of pain catastrophizing (0.57, 95% CI [0.51, 0.63]), pain intensity (0.30, 95% CI [0.24, 0.37]), and pain interference (0.46, 95% CI [0.39, 0.52]). No evidence suggesting differences in this association were found in either the crude or adjusted models (all interaction p-values<0.05).

**Table 2.** Associations between PTSD symptoms and Pain Outcomes in Veterans.

|  | Pain Catastrophizing |  |  | Pain Intensity |  |  | Pain Interference |  |  |
| --- | --- | --- | --- | --- | --- | --- | --- | --- | --- |
| | $\beta$ | 95% CI | Interaction p-value* | $\beta$ | 95% CI | Interaction p-value* | $\beta$ | 95% CI | Interaction p-value* |
| Crude | 0.60 | (0.55, 0.66) | 0.23 | 0.33 | (0.26, 0.39) | 0.67 | 0.47 | (0.41, 0.53) | 0.59 |
| Adjusted | 0.57 | (0.51, 0.63) | 0.22 | 0.30 | (0.24, 0.37) | 0.58 | 0.46 | (0.39, 0.52) | 0.68 |

## Discussion

This study investigated whether the associations between PTSD symptoms and pain catastrophizing, intensity, and interference differed in an approximately equal sample of women and men Veterans. While our bivariate results indicated higher average PTSD symptom scores, pain catastrophizing, pain intensity, and pain interference in women Veterans, the multivariable results did not provide evidence of gender differences in this association. Our results revealed that higher PTSD symptoms had the strongest effect on elevated pain catastrophizing symptoms, followed by pain interference and intensity in both men and women Veterans.

Co-morbidity between PTSD and chronic pain has been well documented in civilian (40) and Veteran samples (4,41). Previous studies assessing the association between PTSD symptoms and pain outcomes in cut-points among Veteran (predominantly men >80%) samples have observed overall higher levels of pain catastrophizing, interference, and intensity (4,5,31,42). The inequity in the gender distribution in these studies left unanswered questions regarding gender differences, given gendered variability in the timing and type of trauma exposure(s) that resulted in PTSD symptoms (26,27). While our results support previous findings, showing robust statistical associations between PTSD symptoms and pain catastrophizing, interference, and intensity, we did not find evidence of gender differences in this association in our gender balanced sample (4,5,41,43).

The present study provides a unique contribution to the existing literature due to the size and approximately equal proportions of men and women Veterans that allowed tests of heterogeneity of effects in this association by gender. The ability to investigate gender differences regarding pain and PTSD in previous Veteran samples has been hampered by poor recruitment and retention of women in VA clinical trials, which typically average only 3-22% women (32,44). The lack of gender differences, may reflect the underlying mechanism of the mutual maintenance model whereby the sensation of pain could trigger PTSD symptoms (intrusions, avoidance, arousal), particularly if the trauma and pain are associated with the same event and this mechanism operates similarly in men and women (28,45). In the case of pain catastrophizing, this may reflect the tendency to magnify the threat value of pain, thereby increasing feelings of helplessness and difficulty inhibiting pain-related thoughts (46). Alternatively, the lack of gender differences in these associations may be in part due to the restriction that all Veterans in the current analysis needed to meet inclusion criteria for entry into the LAMP clinical trial, which may make them more similar compared to the overall Veteran patient population. Eligibility criteria for this clinical trial screened out those with recent diagnosis of a mental disorder (e.g., depression, suicidality), which may have resulted in a clinical sample that is less psychiatrically severe compared to a typical sample of Veterans seeking chronic pain care. These restrictions may obscure gender differences present in more psychiatrically severe patients with comorbid chronic pain conditions.

Elevated levels of pain catastrophizing, intensity, and interference in people with probable PTSD, highlight the need for interdisciplinary care modalities to target both PTSD and chronic pain symptoms. There have been mixed results regarding the efficacy of cognitive-based therapies in reducing comorbid PTSD and pain, with interventions generally demonstrating greater reduction in PTSD symptoms with little change in pain interference and intensity (41,47–49). Mindfulness-based therapies have demonstrated reductions in both PTSD and chronic pain outcomes, indicating their promise as an effective treatment for people with comorbid PTSD and chronic pain (50–54). Finally, Veterans with comorbid chronic pain and PTSD may specifically benefit from integrated and trauma-informed treatment modalities that recognize the mutually reinforcing relationship between PTSD and pain (55). As the VA continues to develop and test multimodal care for Veterans with comorbid trauma and pain conditions, careful attention will be required to evaluate gender-sensitive treatment options (e.g. integrated into women’s clinic, online options) to ensure women Veterans feel comfortable accessing the required care to address these highly co-prevalent conditions (56,57).

This study contributes to the literature by assessing associations between probable PTSD and pain outcomes as well as findings related to gender differences in a Veteran population with highly prevalent PTSD and comorbid chronic pain. Some limitations must be considered when interpreting our results. First, our sample may differ from civilian samples in important ways. For example, compared to civilian samples, Veterans with chronic pain have higher average levels of PTSD symptoms (8). Additionally, we were unable to account for gender differences in the onset of trauma across the life-course (e.g., Adverse Childhood Experience, sexual assault, service-related exposures), where the timing, frequency, and severity may relate to the onset of chronic pain symptoms. Finally, this analysis was conducted on a sample of participants who volunteered to participate in a mindfulness study for chronic pain. Therefore, findings may not generalize to the broader population of Veterans with chronic pain. Despite these limitations, our study has several strengths, including completeness of data, including survey and electronic health record data from a large, diverse, and balanced sample of Veterans with chronic pain. Finally, this uniquely large and gender-balanced study cohort of Veterans allowed for analyses into potential gender differences in pain outcomes by the presence of probable PTSD.

## Conclusion

This study found that PTSD symptoms were associated with elevated levels of pain catastrophizing, pain intensity, and pain interference in women and men Veterans. We did not find any statistically significant gender differences in the associations between PTSD symptoms and pain outcomes. The unique gender composition of this sample created an opportunity to test these differences, addressing a limitation present in other Veteran samples. Our results contribute to research in Veterans aimed at understanding shared underlying mechanisms of this co-morbidity and testing relevant treatments that may concurrently benefit both chronic pain and PTSD.

## Data Availability

These data are the property of the Veterans Administration and contain potentially identifying information about human subjects who use Veterans Health Administration services for their healthcare. Data requests are subject to case-by-case data use agreements and are routed through the Minneapolis VA Research Privacy and Research Compliance officers, the LAMP study PI, and study coordinator. Information regarding the request process is available here: https://www.va.gov/MINNEAPOLISRESEARCH/process/dua.asp. [Used for PLOS ONE submission that requires immediate access to data upon publication.]

## Acknowledgements

The U.S. Army Medical Research Acquisition Activity, 820 Chandler Street, Fort Detrick MD 21702-5014 is the awarding and administering acquisition office. This work was supported by the Assistant Secretary of Defense for Health Affairs endorsed by the Department of Defense, through the Pain Management Collaboratory - Pragmatic Clinical Trials Demonstration Projects under Award No. W81XWH-18-2-0003. Research reported in this publication was supported by Grant Number U24AT009769 from the National Center for Complementary and Integrative Health (NCCIH), and the Office of Behavioral and Social Sciences Research (OBSSR). This material is the result of work supported with resources at the Minneapolis VA Health Care System, Durham VA Health Care System, and VA Greater Los Angeles Healthcare System. For more information about the Collaboratory, visit https://painmanagementcollaboratory.org/.

## References

1. Otis JD, Gregor K, Hardway C, Morrison J, Scioli E, Sanderson K. An examination of the co-morbidity between chronic pain and posttraumatic stress disorder on U.S. Veterans. Psychol Serv. 2010 Aug;7(3):126–35.

2. Shapiro MO, Short NA, Raines AM, Franklin CL, True G, Constans JI. Pain and posttraumatic stress: Associations among women veterans with a history of military sexual trauma. Psychol Trauma Theory Res Pract Policy. 2022;No Pagination Specified-No Pagination Specified.

3. Bair MJ, Outcalt SD, Ang D, Wu J, Yu Z. Pain and Psychological Outcomes Among Iraq and Afghanistan Veterans with Chronic Pain and PTSD: ESCAPE Trial Longitudinal Results. Pain Med. 2020 Jul 1;21(7):1369–76.

4. Benedict TM, Keenan PG, Nitz AJ, Moeller-Bertram T. Post-Traumatic Stress Disorder Symptoms Contribute to Worse Pain and Health Outcomes in Veterans With PTSD Compared to Those Without: A Systematic Review With Meta-Analysis. Mil Med. 2020 Oct;185(9–10):E1481–91.

5. Outcalt SD, Ang DC, Wu J, Sargent C, Yu Z, Bair MJ. Pain experience of Iraq and Afghanistan Veterans with comorbid chronic pain and posttraumatic stress. J Rehabil Res Dev. 2014;51(4):559– 70.

6. Frayne SM, Phibbs CS, Saechao, F. Sourcebook: Women Veterans in the Veterans Health Administration, Volume 4, Longitudinal Trends in Sociodemographics, Utilization, Health Profile, and Geographic Distribution [Internet]. 2018 Feb [cited 2022 Nov 8] p. 144. Available from: https://www.womenshealth.va.gov/docs/WHS_Sourcebook_Vol-IV_508c.pdf

7. Haskell SG, Brandt CA, Krebs EE, Skanderson M, Kerns RD, Goulet JL. Pain among Veterans of Operations Enduring Freedom and Iraqi Freedom: Do Women and Men Differ? Pain Med Malden Mass. 2009;10(7):1167–73.

8. Lehavot K, Katon JG, Chen JA, Fortney JC, Simpson TL. Post-traumatic Stress Disorder by Gender and Veteran Status. Am J Prev Med. 2018 Jan;54(1):e1–9.

9. Nahin RL. Severe Pain in Veterans: The Effect of Age and Sex, and Comparisons With the General Population. J Pain. 2017 Mar;18(3):247–54.

10. How Common is PTSD in Veterans? - PTSD: National Center for PTSD [Internet]. [cited 2023 May 10]. Available from: https://www.ptsd.va.gov/understand/common/common_veterans.asp

11. Andrew R, Derry S, Taylor RS, Straube S, Phillips CJ. The Costs and Consequences of Adequately Managed Chronic Non-Cancer Pain and Chronic Neuropathic Pain. Pain Pract. 2014;14(1):79–94.

12. Gaskin DJ, Richard P. The Economic Costs of Pain in the United States. J Pain. 2012;13(8):715–24.

13. Treede RD, Rief W, Barke A, Aziz Q, Bennett MI, Benoliel R, et al. Chronic pain as a symptom or a disease: the IASP Classification of Chronic Pain for the International Classification of Diseases (ICD-11). Pain. 2019 Jan;160(1):19–27.

14. Voon P, Karamouzian M, Kerr T. Chronic pain and opioid misuse: a review of reviews. Subst Abuse Treat Prev Policy. 2017 Aug 15;12:36.

15. Haskell SG, Heapy A, Reid MC, Papas RK, Kerns RD. The Prevalence and Age-Related Characteristics of Pain in a Sample of Women Veterans Receiving Primary Care. J Womens Health. 2006 Sep;15(7):862–9.

16. Higgins DM, Fenton BT, Driscoll MA, Heapy AA, Kerns RD, Bair MJ, et al. Gender Differences in Demographic and Clinical Correlates among Veterans with Musculoskeletal Disorders. Womens Health Issues. 2017 Jul;27(4):463–70.

17. Teh CF, Kilbourne AM, McCarthy JF, Welsh D, Blow FC. Gender Differences in Health-Related Quality of Life for Veterans With Serious Mental Illness. 2008;59(6).

18. Keefe FJ, Lefebvre JC, Egert JR, Affleck G, Sullivan MJ, Caldwell DS. The relationship of gender to pain, pain behavior, and disability in osteoarthritis patients: the role of catastrophizing. Pain. 2000 Aug 1;87(3):325–34.

19. Keogh E, Boerner KE. Exploring the relationship between male norm beliefs, pain-related beliefs and behaviours: An online questionnaire study. Eur J Pain. 2020;24(2):423–34.

20. Mattocks K, Casares J, Brown A, Bean-Mayberry B, Goldstein KM, Driscoll M, et al. Women Veterans’ Experiences with Perceived Gender Bias in U.S. Department of Veterans Affairs Specialty Care. Womens Health Issues Off Publ Jacobs Inst Womens Health. 2020 Apr;30(2):113–9.

21. Naylor JC, Wagner HR, Johnston C, Elbogen EE, Brancu M, Marx CE, et al. Pain Intensity and Pain Interference in Male and Female Iraq/Afghanistan-era Veterans. Womens Health Issues. 2019 Jun 25;29:S24–31.

22. Sullivan MJL, Tripp DA, Santor D. Gender Differences in Pain and Pain Behavior: The Role of Catastrophizing. Cogn Ther Res. 2000 Feb 1;24(1):121–34.

23. AP Association. Diagnostic and statistical manual of mental disorders: DSM-IV-TR. Am Psychiatr Pub. 2000;157.

24. McCall-Hosenfeld JS, Mukherjee S, Lehman EB. The Prevalence and Correlates of Lifetime Psychiatric Disorders and Trauma Exposures in Urban and Rural Settings: Results from the National Co-morbidity Survey Replication (NCS-R). PLoS ONE. 2014 Nov 7;9(11):e112416.

25. Street AE, Vogt D, Dutra L. A new generation of women veterans: Stressors faced by women deployed to Iraq and Afghanistan. Clin Psychol Rev. 2009 Dec 1;29(8):685–94.

26. Vogt D, Vaughn R, Glickman ME, Schultz M, Drainoni ML, Elwy R, et al. Gender differences in combat-related stressors and their association with postdeployment mental health in a nationally representative sample of U.S. OEF/OIF veterans. J Abnorm Psychol. 2011 Nov;120(4):797–806.

27. Driscoll MA, Higgins DM, Seng EK, Buta E, Goulet JL, Heapy AA, et al. Trauma, Social Support, Family Conflict, and Chronic Pain in Recent Service Veterans: Does Gender Matter? Pain Med Malden Mass. 2015;16(6):1101–11.

28. Asmundson GJ, Coons MJ, Taylor S, Katz J. PTSD and the Experience of Pain: Research and Clinical Implications of Shared Vulnerability and Mutual Maintenance Models. Can J Psychiatry. 2002 Dec 1;47(10):930–7.

29. Sharp TJ, Harvey AG. Chronic pain and posttraumatic stress disorder: mutual maintenance? Clin Psychol Rev. 2001 Aug 1;21(6):857–77.

30. McAndrew LM, Lu SE, Phillips LA, Maestro K, Quigley KS. Mutual maintenance of PTSD and physical symptoms for Veterans returning from deployment. Eur J Psychotraumatology. 2019 Dec 31;10(1):1608717.

31. Outcalt SD, Kroenke K, Krebs EE, Chumbler NR, Wu J, Yu Z, et al. Chronic pain and comorbid mental health conditions: independent associations of posttraumatic stress disorder and depression with pain, disability, and quality of life. J Behav Med. 2015 Jun;38(3):535–43.

32. Danan ER, Ullman K, Klap RS, Yano EM, Krebs EE. Evidence Map: Reporting of Results by Sex or Gender in Randomized, Controlled Trials with Women Veteran Participants (2008 to 2018). Womens Health Issues Off Publ Jacobs Inst Womens Health. 2019 Jun 25;29 Suppl 1(Suppl 1):S112–20.

33. Burgess DJ, Evans R, Allen KD, Bangerter, A, Bronfort G, Cross LJ, et al. Learning to Apply Mindfulness to Pain (LAMP): Design for a Pragmatic Clinical Trial of Two Mindfulness-Based Interventions for Chronic Pain. Pain Med. 2020 Dec 12;21(Supplement_2):S29–36.

34. Bovin MJ, Marx BP, Weathers FW, Gallagher MW, Rodriguez P, Schnurr PP, et al. Psychometric properties of the PTSD Checklist for Diagnostic and Statistical Manual of Mental Disorders-Fifth Edition (PCL-5) in veterans. Psychol Assess. 2016 Nov;28(11):1379–91.

35. Using the PTSD Checklist for DSM-5 (PCL-5).

36. Sullivan MJL, Bishop SR, Pivik J. The Pain Catastrophizing Scale: Development and validation. Psychol Assess. 1995 Dec;7(4):524–32.

37. Cleeland CS, Ryan KM. Pain assessment: Global use of the Brief Pain Inventory. Ann Acad Med Singap. 1994;23(2):129–38.

38. Keller S, Bann CM, Dodd SL, Schein J, Mendoza TR, Cleeland CS. Validity of the Brief Pain Inventory for Use in Documenting the Outcomes of Patients With Noncancer Pain: Clin J Pain. 2004;20(5):309–18.

39. Goldsmith ES, Taylor BC, Greer N, Murdoch M, MacDonald R, McKenzie L, et al. Focused Evidence Review: Psychometric Properties of Patient-Reported Outcome Measures for Chronic Musculoskeletal Pain. J Gen Intern Med. 2018 May;33(Suppl 1):61–70.

40. Slawek DE, Syed M, Cunningham CO, Zhang C, Ross J, Herman M, et al. Pain catastrophizing and mental health phenotypes in adults with refractory chronic pain: A latent class analysis. J Psychiatr Res. 2021 Dec 2;145:102–10.

41. Goldstein E, McDonnell C, Atchley R, Dorado K, Bedford C, Brown RL, et al. The Impact of Psychological Interventions on Posttraumatic Stress Disorder and Pain Symptoms: A Systematic Review and Meta-Analysis. Clin J Pain. 2019 Aug;35(8):703–12.

42. Alschuler KN, Otis JD. Coping strategies and beliefs about pain in veterans with comorbid chronic pain and significant levels of posttraumatic stress disorder symptoms. Eur J Pain Lond Engl. 2012 Feb;16(2):312–9.

43. Asmundson GJG, Wright KD, Stein MB. Pain and PTSD symptoms in female veterans. Eur J Pain. 2004 Aug 1;8(4):345–50.

44. Goldstein KM, Duan-Porter W, Alkon A, Olsen MK, Voils CI, Hastings SN. Enrollment and Retention of Men and Women in Health Services Research and Development Trials. Womens Health Issues Off Publ Jacobs Inst Womens Health. 2019 Jun 25;29 Suppl 1(Suppl 1):S121–30.

45. Brennstuhl MJ, Tarquinio C, Montel S. Chronic Pain and PTSD: Evolving Views on Their Comorbidity: Chronic Pain and PTSD: Evolving Views on Their Comorbidity. Perspect Psychiatr Care. 2015 Oct;51(4):295–304.

46. Quartana PJ, Campbell CM, Edwards RR. Pain catastrophizing: a critical review. Expert Rev Neurother. 2009 May;9(5):745–58.

47. Dunne RL, Kenardy J, Sterling M. A Randomized Controlled Trial of Cognitive-behavioral Therapy for the Treatment of PTSD in the Context of Chronic Whiplash. Clin J Pain. 2012 Nov;28(9):755–65.

48. Beck JG, Coffey SF, Foy DW, Keane TM, Blanchard EB. Group Cognitive Behavior Therapy for Chronic Posttraumatic Stress Disorder: An Initial Randomized Pilot Study. Behav Ther. 2009 Mar;40(1):82–92.

49. Jull G, Kenardy J, Hendrikz J, Cohen M, Sterling M. Management of acute whiplash: A randomized controlled trial of multidisciplinary stratified treatments. Pain. 2013 Sep;154(9):1798–806.

50. Plagge JM, Lu MW, Lovejoy TI, Karl AI, Dobscha SK. Treatment of Comorbid Pain and PTSD in Returning Veterans: A Collaborative Approach Utilizing Behavioral Activation. Pain Med. 2013 Aug 1;14(8):1164–72.

51. Okvat HA, Davis MC, Mistretta EG, Mardian AS. Mindfulness-based training for women veterans with chronic pain: A retrospective study. Psychol Serv. 2022;19(Suppl 1):106–19.

52. King AP, Block SR, Sripada RK, Rauch SAM, Porter KE, Favorite TK, et al. A Pilot Study of Mindfulness-Based Exposure Therapy in OEF/OIF Combat Veterans with PTSD: Altered Medial Frontal Cortex and Amygdala Responses in Social–Emotional Processing. Front Psychiatry [Internet]. 2016 Sep 20 [cited 2023 Jun 16];7. Available from: 10.3389/fpsyt.2016.00154/abstract

53. Schure MB, Simpson TL, Martinez M, Sayre G, Kearney DJ. Mindfulness-Based Processes of Healing for Veterans with Post-Traumatic Stress Disorder. J Altern Complement Med. 2018 Nov;24(11):1063–8.

54. Marchand WR, Sandoval K, Lackner R, Parker SC, Herrmann T, Yabko B, et al. Mindfulness-based interventions for military veterans: A systematic review and analysis of the literature. Complement Ther Clin Pract. 2021 Feb;42:101274.

55. Pebole M, Gobin RL, Hall KS. Trauma-informed exercise for women survivors of sexual violence. Transl Behav Med. 2021 Mar 16;11(2):686–91.

56. Fenwick KM, Golden RE, Frayne SM, Hamilton AB, Yano EM, Carney DV, et al. Women Veterans’ Experiences of Harassment and Perceptions of Veterans Affairs Health Care Settings During a National Anti-Harassment Campaign. Womens Health Issues. 2021 Nov;31(6):567–75.

57. Kelly U, Haywood T, Segell E, Higgins M. Trauma-Sensitive Yoga for Post-Traumatic Stress Disorder in Women Veterans who Experienced Military Sexual Trauma: Interim Results from a Randomized Controlled Trial. J Altern Complement Med. 2021 Mar 1;27(S1):S-45-S-59.

